# Trends in TSH, free T4, and anti-thyroid peroxidase and treatment status: Canadian Health Measures Survey 2012 to 2015

**DOI:** 10.1101/2022.04.21.22274148

**Authors:** Yi-Sheng Chao

**Affiliations:** Independent researcher

**Keywords:** Canadian Health Measures Survey (CHMS), trends, thyroid hormone, thyroid-stimulating hormone, treatment status

## Abstract

**Background:** Previous studies in Canada focused on the prevalence of thyroid conditions have not reported on the levels of the thyroid-stimulating hormone (TSH) and thyroid hormones. To address this issue, the present study describes the trends in TSH, free T4, and anti-thyroid peroxidase and their treatment status for the patients who have clinically high or low levels.

**Methods:** We used data from the Canadian Health Measures Survey (CHMS) cycles 3 and 4 conducted between 2012 and 2015. The thyroid measures studied were TSH, free T4, and anti-thyroid peroxidase. We used clinical reference ranges to identify abnormality in these measures. We labelled abnormality in these measures as *treated* if relevant conditions were diagnosed or a disease-specific prescription was reported. *Untreated* individuals were those with an abnormality in thyroid measures without any medication use or relevant diagnoses. We presented the trends of thyroid measures in mean values and ratios, compared to the values first measured.

**Results:** The levels of TSH, free T4, and anti-peroxidase in cycle 4 were not significantly different from those in cycle 3. The proportions of Canadians with clinically high levels of free T4, anti-thyroid peroxidase, and TSH were 0.03 to 0.017, 0.005 to 0.005, and 0.30 to 0.43 for cycles 3 to 4, respectively. The proportions of Canadians with clinically low levels of TSH and free T4 were 0.02 to 0.021 and 0.18 to 0.11 for cycles 3 to 4, respectively. The change in the proportions of treatment statuses varied across the thyroid measures of the Canadians studied.

**Conclusion:** This descriptive study demonstrates the trends in TSH, free T4, and anti-thyroid peroxidase; their distributions in the population; and the proportions of Canadians with clinically high or low levels. We believe the information on the treatment status of those with uncontrolled high levels can be used to design patient screening programs.

## Background

It was estimated that in 2008 and 2009, 10.4% of Canadians aged 45 years or older had thyroid conditions (unspecified).[1, 2] Not all patients present symptoms and are treated.[1] However, thyroid conditions left untreated can lead to adverse cardiac outcomes, such as hypertension and dyslipidemia.[1] Asymptomatic patients can be identified through population screening. The screening of thyroid conditions often is done by measuring the levels of blood thyroid-stimulating hormone (TSH).[1] In clinics, free thyroxine (T4) and anti-thyroid peroxidase are two of the thyroid measures tested to determine a differential diagnosis of hyper-or hypothyroidism.[3]

Certain treatment options are available for asymptomatic thyroid conditions, such as levothyroxine for screen-detected hypothyroidism.[1] However, concerns have be raised with respect to the population screening for thyroid conditions. First, levels of TSH may fluctuate. For example, a 5-year cohort study found that abnormal levels of TSH in some primary care patients returned to normal levels.[4] Second, no strong evidence exists to support treatment for asymptomatic patients. A systematic review by the Canadian Task Force on Preventive Health Care found a lack of statistical significance in the differences in mortality, weight change, and harms between the treated and untreated screen-detected thyroid patients.[1] Therefore, screening the thyroid conditions of the general public is not recommended, with the exception of those individuals with previous thyroid conditions or those who have received thyroid interventions.[1] Last, overdiagnosis is a concern that may occur when a condition is detected that would not otherwise have been diagnosed in an individual’s lifetime.[5] Globally, in developed and developing countries, a trend in thyroid cancer overdiagnosis is increasing.[5] For example, in the Republic of Korea, a 15-fold increase occurred in the diagnosis of thyroid cancer between 1993 and 2011, even though the thyroid cancer mortality rate remained similar to the 1993 level throughout the period.[6] Thus, a trend of overdiagnosis of thyroid cancer in the Republic of Korea is likely.[6]

In Canada, thyroid cancer is not targeted by cancer screening programs[7], and screening for thyroid conditions is not recommended.[1] It is unclear whether the lack of support for screening is the reason why studies reporting the statistics on TSH and other thyroid measures are few. Currently, little information is available as to whether an overdiagnosis or undertreatment of thyroid conditions is occurring. Previous studies have used a trend analysis of blood pressure and environmental chemicals to demonstrate their distributions in the populations and to detect significant changes in the levels of biomarkers over time.[8-10] The trends in, and the treatment status of, TSH and thyroid hormones may be useful for understanding how thyroid conditions are clinically managed in Canada. Therefore, this descriptive study conducts a trend analysis of TSH, free T4, and anti-thyroid peroxidase—using the thyroid measures available in a national survey—to detect whether significant changes in their levels are occurring in Canadians, and to estimate the treatment statuses of these thyroid measures.

## Methods

We used data from the Canadian Health Measures Survey (CHMS) cycles 3 and 4 implemented in 2012 to 2013 and 2014 to 2015, respectively.[8-10] For each of these cycle, more than 5,000 non-institutionalized Canadians, aged 3 to 79 years were sampled (for details about inclusion and exclusion criteria, see previous publications).[11, 12] statistical weights were applied to the subjects to estimate the overall distributions and levels of selected measures for more than 30 million non-institutionalized Canadians.[13] Three thyroid measures were available for the CHMS cycles 3 and 4: free T4 (pmol/l), anti-thyroid peroxidase (U/ml), and TSH (mIU/l).[14]

### Abnormality identification for blood measures

We assessed TSH, free T4, and anti-thyroid peroxidase for abnormality, values higher or lower than upper or lower reference limits, respectively. The reference ranges for TSH were 0.4 to 4.8 mIU/l for adults and 1.7 to 9.1 mIU/l for those aged 21 weeks to 20 years.[15] The reference ranges for free T4 were 13 to 27 pmol/l.[15] The upper limit of the anti-thyroid peroxidase reference range was 35 U/ml.[16]

### Treatment status identification

We defined the *treatment status* according to the use of medication and the chronic condition diagnoses reported by participants.[17-20] We estimated the treatment status in four categories: treated, probably treated, potentially treated, and untreated (see Table 1). To avoid over-identification, we defined *untreated individuals* as those who had not reported the use of any medications or any related chronic conditions defined by the CHMS (any chronic conditions, cardiovascular disease, high blood pressure, abnormal lipid profile, stroke, kidney disease, liver disease, thyroid disease, diabetes, cancer, eating disorder, or chronic obstructive pulmonary disease). We regarded individuals as *treated* only if they were diagnosed with thyroid disease or who were taking thyroid prescription drugs.

**Table 1.** The conditions and medications used to define the treatment status of TSH, free T4, and anti-thyroid peroxidase in the Canadian Health Measures Survey, cycles 3 to 4 implemented in 2012 to 2013 and 2014 to 2015, respectively.

| Disease categories | CHMS variables | CHMS concept | Medication | ATC codes | CHMS variables | CHMS concepts | Units |
| --- | --- | --- | --- | --- | --- | --- | --- |
| Thyroid condition | ccc_82 | Has a thyroid condition | Thyroid | First and second level: G03 | lab_ft4 | Free thyroxine | pmol/l |
|  |  |  |  |  | lab_tsh | Thyroid stimulating hormone (TSH) or thyrotropin | pmol/l |
|  |  |  |  |  | lab_tpo | Anti-thyroid peroxidase | IU/ml |

ATC = Anatomical Therapeutic Chemical; CHMS = Canadian Health Measures Survey; TSH = thyroid stimulating hormone. There are five levels in the ATC classification. The first level is represented by the first character of the codes. The second levels are the two digits following the first character.

Individuals were *probably treated* if they were diagnosed with the above-mentioned chronic conditions that might affect clinical measures or were taking any prescription drugs. Individuals were regarded as *potentially treated* if they reported any chronic conditions or were taking any prescription or over-the-counter drugs.

We reported *medication use* based on the Anatomical Therapeutic Chemical (ATC) classification system.[9, 21] *Medication* in the CHMS data was labelled as prescription, over-the-counter drugs, and health products.[9] Over-the-counter drugs and health products were regarded as *non-prescription*.[9] We extracted the ATC codes that represented prescription or over-the-counter drugs from the medication file of each cycle. In Table 1, the first and second levels of the ATC codes that represented thyroid medications were indicated as “G03.”

### Data analysis

We conducted a trend analysis using all the variables available from the CHMS.[8, 9, 22] First, we summarized these variables using basic characteristics, such as maximal and minimal values, and we linked the survey and bootstrap weights to them.[22] Then, we identified the missing values and the values beyond the detection levels for all the variables.[10] We recorded basic statistics, such as the weighted mean, and the 25th and 75th percentiles.[10] We introduced clinical reference ranges to identify Canadians with abnormal levels of thyroid measures. We labelled four treatment statuses based on chronic conditions and medication use. We used the CHMS cycles as the proxy measure for time points in our trend analysis. Using ratios, we compared the distributions of TSH, free T4, and anti-thyroid peroxidase in CHMS cycle 2 (2012 to 2013) to their distributions in CHMS cycle 3 (2014 to 2015).[9, 10, 23] We described the trends with ratios with 95% confidence intervals (CIs). The 95% CIs of the ratios in the subsequent cycles covering 1 indicated insignificant changes in the levels of thyroid measures after cycle 3. We conducted the data analysis with R (v3.20)[24] and RStudio (v0.98.113).[25]

## Results

The population characteristics in the same period were published.[9, 10] In summary, throughout the CHMS cycles 3 to 4, around 30 million Canadians were estimated and almost half of them were female.[9, 10] The mean ages of the two cycles were 39.00 and 39.30 years.[8-10] The trends of free T4, anti-peroxidase, and TSH of cycles 3 and 4 are represented in Tables 2, 3, and 4. The levels of free T4 were all within the detection limits. The proportions of individuals reaching the lower detection limits were 0.39, 0.30, and less than 0.0002 for anti-thyroid peroxidase in cycles 3 and 4 and TSH in cycle 3, respectively. Since the lower reference range for anti-thyroid peroxidase was not defined, and the proportion of TSH lower than the detection levels was small, the proportions of thyroid measures lower than the detection levels did not influence the subsequent analysis. Based on the ratios in cycle 4, the levels of TSH, free T4, and anti-thyroid peroxidase were not significantly different from those levels in cycle 3 (ratio 95% CIs covering 1 for all).

**Table 2.**
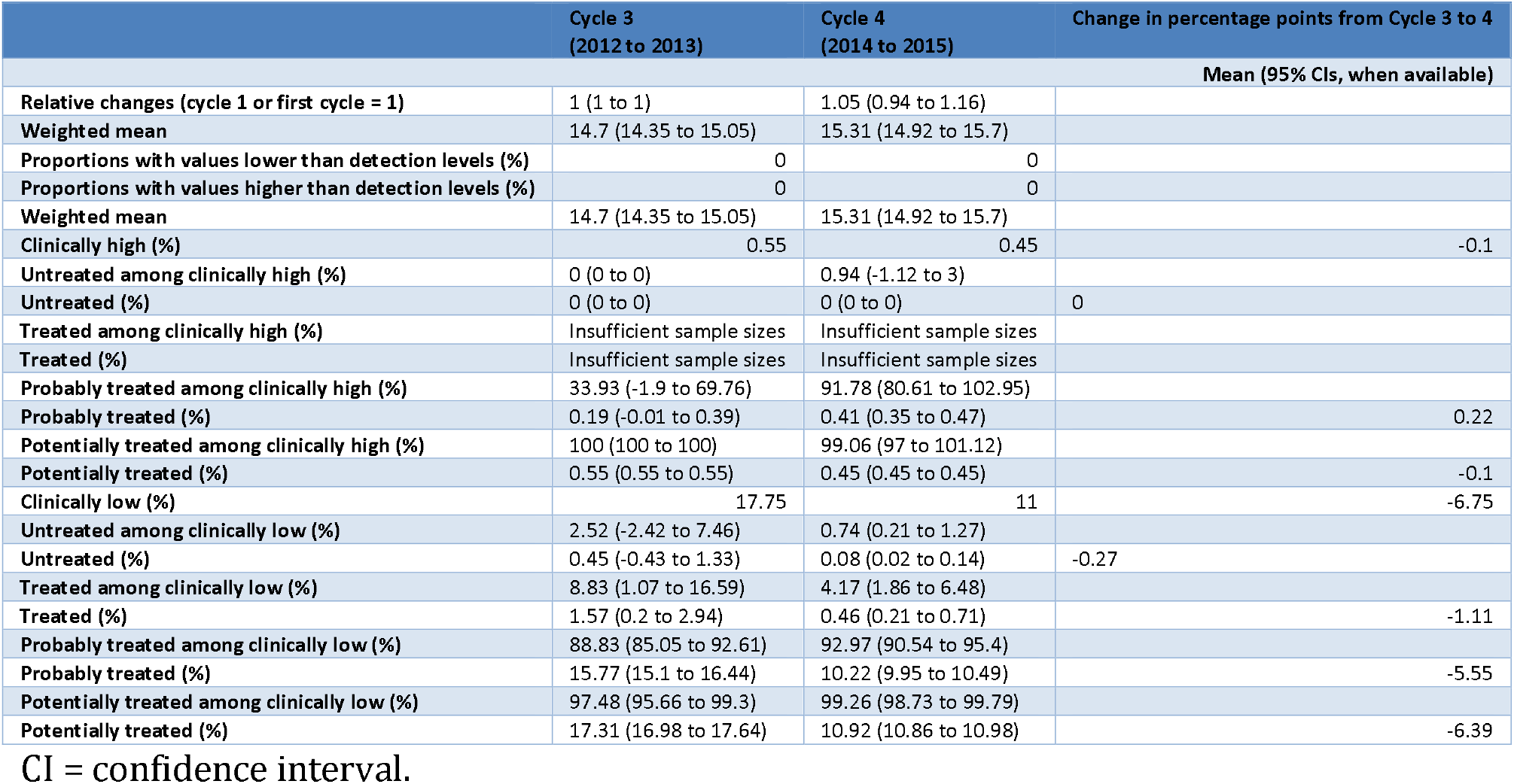
Trends in the Levels of Free T4 (thyroxine, pmol/l) and the Treatment Statuses in the Canadian Health Measures Survey Cycles 3 and 4.

**Table 3.**
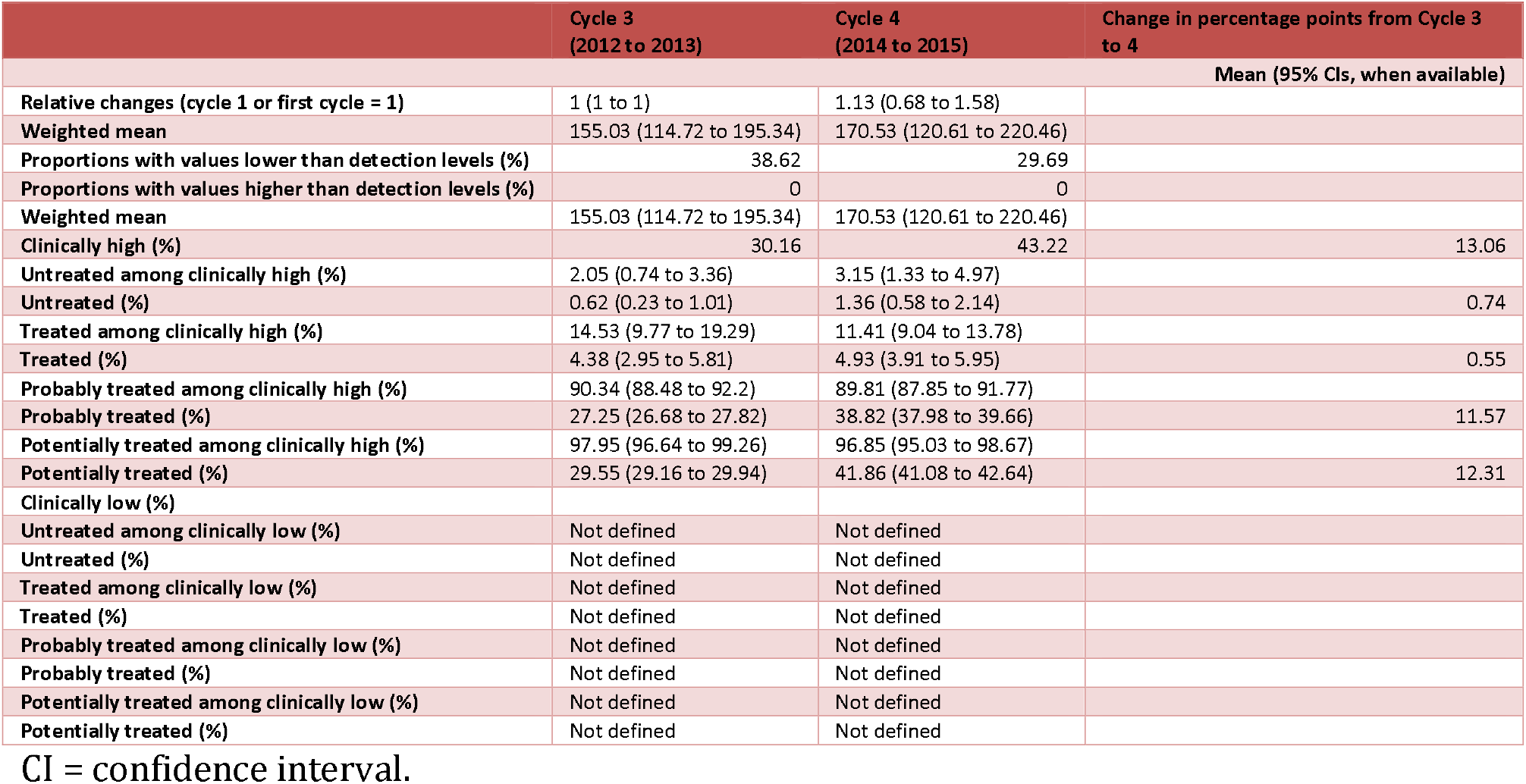
Trends in the Levels of Antithyroid Peroxidase (U/ml) and the Treatment Statuses in the Canadian Health Measures Survey Cycles 3 and 4.

|  | Cycle 3<br>(2012 to 2013) | Cycle 4<br>(2014 to 2015) | Change in percentage points from Cycle 3<br>to 4 |
| --- | --- | --- | --- |
|  | Mean (95% CIs, when available) |  |  |
| Relative changes (cycle 1 or first cycle = 1) | 1 (1 to 1) | 1.13 (0.68 to 1.58) |  |
| Weighted mean | 155.03 (114.72 to 195.34) | 170.53 (120.61 to 220.46) |  |
| Proportions with values lower than detection levels (%) | 38.62 | 29.69 |  |
| Proportions with values higher than detection levels (%) | 0 | 0 |  |
| Weighted mean | 155.03 (114.72 to 195.34) | 170.53 (120.61 to 220.46) |  |
| Clinically high (%) | 30.16 | 43.22 | 13.06 |
| Untreated among clinically high (%) | 2.05 (0.74 to 3.36) | 3.15 (1.33 to 4.97) |  |
| Untreated (%) | 0.62 (0.23 to 1.01) | 1.36 (0.58 to 2.14) | 0.74 |
| Treated among clinically high (%) | 14.53 (9.77 to 19.29) | 11.41 (9.04 to 13.78) |  |
| Treated (%) | 4.38 (2.95 to 5.81) | 4.93 (3.91 to 5.95) | 0.55 |
| Probably treated among clinically high (%) | 90.34 (88.48 to 92.2) | 89.81 (87.85 to 91.77) |  |
| Probably treated (%) | 27.25 (26.68 to 27.82) | 38.82 (37.98 to 39.66) | 11.57 |
| Potentially treated among clinically high (%) | 97.95 (96.64 to 99.26) | 96.85 (95.03 to 98.67) |  |
| Potentially treated (%) | 29.55 (29.16 to 29.94) | 41.86 (41.08 to 42.64) | 12.31 |
| Clinically low (%) |  |  |  |
| Untreated among clinically low (%) | Not defined | Not defined |  |
| Untreated (%) | Not defined | Not defined |  |
| Treated among clinically low (%) | Not defined | Not defined |  |
| Treated (%) | Not defined | Not defined |  |
| Probably treated among clinically low (%) | Not defined | Not defined |  |
| Probably treated (%) | Not defined | Not defined |  |
| Potentially treated among clinically low (%) | Not defined | Not defined |  |
| Potentially treated (%) | Not defined | Not defined |  |

CI = confidence interval.

**Table 4.**
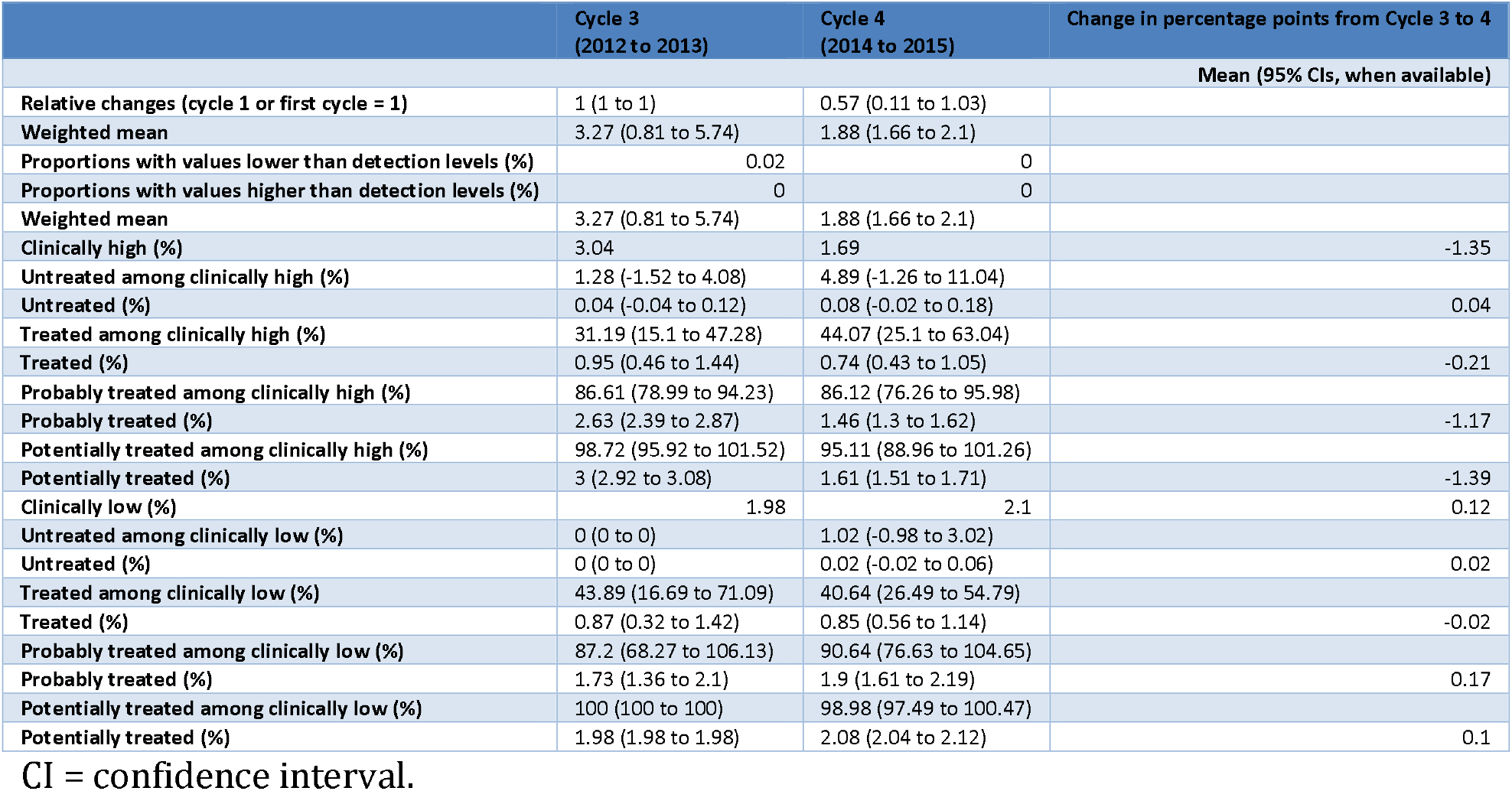
Trends in the Levels of Thyroid Stimulating Hormone (mIU/l) and the Treatment Statuses in the Canadian Health Measures Survey Cycles 3 and 4.

### Proportions of abnormality in blood measures

The proportions of Canadians with levels higher or lower than the reference ranges differed for TSH, free T4, and anti-thyroid peroxidase. There was an increase in the proportions of Canadians with high levels of anti-thyroid peroxidase and low levels of TSH. There was a decrease in the proportions of Canadians with high levels of free T4 and TSH, and low levels of free T4. The proportions of Canadians with clinically high levels of free T4, anti-thyroid peroxidase, and TSH were 0.005 to 0.005, 0.30 to 0.43, and 0.03 to 0.017 for cycles 3 to 4, respectively. The proportions of Canadians with clinically low levels of free T4 and TSH were 0.18 to 0.11 and 0.02 to 0.021 for cycles 3 to 4, respectively. The lower reference limit for anti-thyroid peroxidase was not defined, and the proportion with clinically low levels was not available.

### Treatment status for clinically high levels

The four estimated treatment status varied for TSH, free T4, and anti-thyroid peroxidase. Due to an insufficient number of unweighted sample sizes, the proportions of Canadians being treated with high levels of free T4 were not reported in Table 2. The proportions of Canadians who were untreated—based on drug use and self-reported conditions—were all less than 5% of Canadians with high levels of free T4, anti-thyroid peroxidase, and TSH: 0 to 0.009, 0.02 to 0.03, and 0.01 to 0.049 for cycle 3 and 4, respectively.

The proportions of Canadians treated with high levels of TSH and anti-thyroid peroxidase were 0.31 to 0.44 and 0.15 to 0.11 for cycles 3 to 4, respectively; whereas sufficient sample sizes were unavailable for estimating this proportion for free T4.

The present study uniquely identified two treatment statuses—*probably treated* or *potentially treated*. Among those with high levels of free T4, anti-thyroid peroxidase, and TSH, the proportions of being probably treated were high: 0.34 to 0.92, 0.90 to 0.90, and 0.87 to 0.86 for cycles 3 to 4, respectively. Among Canadians with high levels of free T4, anti-thyroid peroxidase, and TSH, the proportions of being probably treated were high: 1 to 0.99, 0.98 to 0.97, and 0.99 to 0.95 for cycles 3 to 4, respectively.

### Treatment status for clinically low levels

The treatment status for clinically low levels of anti-thyroid peroxidase was not defined. The proportions of Canadians with low levels of free T4 and TSH were 0.18 to 0.11 and 0.020 to 0.021 for cycles 3 to 4, respectively. The proportions of *untreated* Canadians with low levels of free T4 and TSH were 0.025 to 0.007 and 0 to 0.01 for cycles 3 to 4, respectively. The proportions of Canadians being *treated* who had low levels of free T4 and TSH were 0.09 to 0.04 and 0.44 to 0.41 for cycles 3 to 4, respectively.

The proportions of Canadians being *probably treated* were high for free T4 and TSH. Thus, the proportions of Canadians with low levels of free T4 and TSH being probably treated were 0.89 to 0.93 and 0.87 to 0.91 for cycles 3 to 4, respectively. The proportions of Canadians being *potentially treated* were almost 1 for free T4 and TSH. Thus, the proportions of Canadians with low levels of free T4 and TSH being potentially treated were 0.97 to 0.99 and 1 to 0.99 for cycles 3 to 4, respectively.

## Discussion

Surprisingly, based on our findings, little attention has been paid to the trends in thyroid measures, their distributions in the population, and the clinically low levels of thyroid measures. Building on previous studies,[26] we did not limit the scope of our study to subjects with clinically high levels.

The levels of TSH, free T4, and anti-thyroid peroxidase were stable throughout CHMS cycles 3 to 4. According to the CHMS surveys, Canadians have high proportions of abnormal (high or low) levels of anti-thyroid peroxidase and free T4 (more than 11%). The proportions of Canadians with abnormal levels of TSH, a screening target, were 5% or less. Also, the proportions of Canadians with high levels of anti-thyroid peroxidase and low levels of TSH may have increased, and the proportions of Canadians with high levels of free T4 and TSH and low levels of free T4 may have decreased.

High proportions of Canadians with abnormal levels of TSH, free T4, and anti-thyroid peroxidase were probably treated, taking any prescriptions, or diagnosed with thyroid disease or other chronic conditions, which may suggest a high health care coverage for Canadians with abnormal levels of TSH, free T4, and anti-thyroid peroxidase.

A previous study using CHMS cycle 3 data with the same population and a slightly different reference range (0.55 to 4.78 mIU/l, compared to 0.4 to 4.8 mIU/l of the present study) found that the proportions of Canadians with clinically high, normal, and low levels of TSH in cycle 3 were 3.33%, 93.15%, and 3.52%, respectively, compared to 3.04%, 94.98%, and 1.98% of the present study.[27] The similarity of these estimates suggests that the analysis of the present study is accurate.

Since Canada lacks a comprehensive reporting of thyroid conditions, the prevalence of thyroid conditions in Canada is based on sporadic reports. Between 2008 and 2009, the prevalence of thyroid conditions in the Canadian population was 10.4% among those aged 45 years and over.[2] The estimated prevalence of thyroid condition was 5.67% and 6.85% among Canadians aged 3 and over in 2009 to 2011 and 2012 to 2013, respectively.[27] In the US, the prevalence of thyroid conditions was estimated to be 16.18% in women and 3.06% in men, reported by participants interviewed between 1999 and 2006.[28] This study and others have shown that some of the patients with abnormal levels of TSH remain undiagnosed.[28, 29]

### Treatment and clinically abnormal levels

One feature of the present study is its four treatment statuses indicating the intensity of medical interventions that Canadians have received. Moreover, the proportions of the four treatment statuses that identified TSH, free T4, and anti-thyroid peroxidase were different. The first finding based on treatment statuses is that thyroid medications are rarely used by Canadians with high levels of free T4. An increase in the levels of free T4 has been linked to increased risks of developing any solid cancer, lung cancer, and breast cancer, regardless of the TSH levels.[30] This outcome may be due to the activation of ontogenetic pathways and tumor-growth-related genes.[30] The second finding is that the majority of Canadians with abnormal levels of TSH, free T4, or anti-thyroid peroxidase were probably treated (were taking prescriptions or diagnosed with chronic conditions), more than 86%, except free T4 in cycle 3 (34%). Whether this is due to an increase in the treatment coverage, changes to clinical practice, or differences in sampling methods needs to be confirmed with future CHMS cycles.

The third finding based on treatment statuses is that although individuals may be treated, the levels of thyroid measures remain abnormal, i.e., inadequately controlled. The proportions of individuals with inadequately controlled levels of lipid profile in the general public have been used to assess the performance of health care systems.[18] TSH, free T4, and total T3 are important measures to follow-up with patients who have a thyroid condition.[31, 32] Our estimates of the proportions of uncontrolled high or low levels of TSH are more than 31% or 40% among those with high or low levels of TSH, respectively. The proportions of uncontrolled high or low levels of free T4 are less than 1% or more than 4% among those with high or low levels of free T4, respectively. We did not find similar statistics reported by other researchers. Whether these present study statistics are useful for assessing the efficacy of thyroid condition intervention remains to be investigated.

The fourth finding based on treatment statuses is that the association of medical interventions with the proportions of clinically high or low levels of TSH, free T4, and anti-thyroid peroxidase varies. Various factors can modify treatment efficacy, such as intervention duration, treatment doses, frequency,[33], and compliance to medical interventions.[20, 34, 35] It remains unclear whether the proportions of inadequately controlled abnormality of thyroid measures is a useful indicator for assessing intervention adequacy for those patients with abnormal levels of TSH, free T4, or anti-thyroid peroxidase.

Last, opportunities may exist to intervene based on the association of treatment statuses and abnormality in blood measures. The differences in the proportions of *being treated* and *potentially treated* among Canadians with clinically high or low levels are more than 50% for TSH, free T4, and anti-thyroid peroxidase in cycles 3 and 4. The difference between being probably treated and potentially treated is taking over-the-counter medications, which suggests that most Canadians with abnormal levels of TSH, free T4, or anti-thyroid peroxidase were taking some medication. With respect to TSH, this suggests that more than 94% of Canadians with clinically high or low levels may be targeted for interventions through their regular access to medication. Several pharmacist-led initiatives have been designed to screen conditions for treatment, for example, sexually transmitted infections,[36] sleep apnea,[37] asthma, obesity, and hypertension.[38] Also, pediatric patients have been identified by pharmacist-led screening programs.[38] Our results may help to refine the directions and magnitudes that health authorities can use to design population screening programs when necessary.

### Overdiagnosis of thyroid conditions?

Korean researchers used a 15-fold increase in thyroid cancer diagnosis prevalence and stable thyroid cancer mortality to demonstrate the possible extent of overdiagnosis of thyroid cancer.[6] The present study was not designed to directly assess an overdiagnosis or undertreatment of a thyroid condition, but the trends in, and the treatment statuses of, thyroid measures can be used as the foundation for assessing these issues. Thus, the present study findings can be informative for planning an investigation of the overdiagnosis or undertreatment of thyroid conditions. First, it may not be adequate to assess the trends in thyroid treatment solely based on the use of a thyroid prescription (ATC codes beginning with G03) because the use rate is low among those with abnormal levels of TSH. Second, the levels of TSH, free T4, and anti-thyroid peroxidase remain similar throughout the CHMS cycles 3 and 4. These trends provide important baseline data for future monitoring. Whether the trends in TSH, free T4, and anti-thyroid peroxidase may be changed or altered by other factors can be studied with such data.

Last, the assessment of overdiagnosis is difficult with information based only on blood measures and treatment status at two time points. A long-term follow-up and major health outcomes, such as mortality, will be needed for assessing an overdiagnosis.[6] At the same time, the measures of the CHMS are subject to changes in medical practices and sampling frames, or to measurement errors.[8] Without obvious signs in CHMS cycles 3 and 4, this issue needs to be monitored in the future.

### Sick population versus sick individuals

The results of the present study show that large proportions of Canadians with clinically high or low levels of anti-thyroid peroxidase and free T4 may highlight the shortcomings of current health care that focus more on sick individuals than the signs of increasing risks to thyroid health in the general population.[39] Previous Canadian studies have reported on only the proportions of patients with clinically high or low levels of TSH.[27] With respect to the Canadian population, the distributions of TSH, free T4, and anti-thyroid peroxidase have not been well studied, let alone the trends of their distributions. Our research group has been developing tools to analyze trends in national surveys[8-10] and to demonstrate the importance of trends in blood measures and environmental chemicals.[8-10] If the increasing risks of thyroid conditions are limited only to certain subgroups, preventive strategies should monitor and focus on individuals with such characteristics, i.e., the high-risk strategies described by Geoffrey Rose.[39] However, the increase in risks applies to the general population. This strategy will require policies that do not focus on certain individuals, i.e., the population strategies described by Geoffrey Rose.[39] We think the results of this trend analysis provide a good foundation for a discussion on how to reduce the risk for Canadians to develop long-term consequences of untreated or undertreated thyroid conditions.

### Limitations

Fluctuations of large magnitudes in other blood measures or environmental chemicals have occurred throughout CHMS cycles 1 to 4.[8] Besides the changes in the distributions of blood measures across time, other factors may lead to different levels of blood measures or various proportions of treatment statuses over time, such as differences in sampling frames, quantification methods, or precisions; and changes in detection levels.[8] Moreover, a lack of overall significant trends in population levels may not mean a lack of significant changes in blood measure levels among subpopulations. The CHMS data are not designed to track or follow-up with individuals with specific characteristics or smaller geographic areas[11-13], and so it may not be adequate to conclude that significant trends in all subpopulations are lacking.

## Conclusion

Based on our findings, little attention has been paid to trends in TSH, free T4, and anti-thyroid peroxidase; their distributions in the population; and clinically low levels of these measures. Based on the ratios from cycles 3 to 4, significant changes are lacking in the levels of TSH, free T4, and anti-thyroid peroxidase. There are more than 30% and 11% of Canadians with clinically high or low levels (undetected or uncontrolled by medication) of anti-thyroid peroxidase and free T4, respectively. The proportions of the four treatment statuses, which represent the different treatment intensities that patients may have received, are different for TSH, free T4, and anti-thyroid peroxidase. Our results show that the large proportions of Canadians with clinically high or low levels of TSH, free T4, or anti-thyroid peroxidase may highlight the shortcomings of current health care that focus more on sick individuals than the signs of increasing risks to the thyroid health of the general population. We plan to research strategies to screen undiagnosed patients, and the potential causes of the high proportions of uncontrolled thyroid measures.

## Data Availability

The CHMS data are not publicly available. It is against the Statistics Act of Canada to release the CHMS data. The collection of the CHMS data has been approved by the ethics committee within the governments of Canada. The CHMS data have been de-identified and maintained by Statistics Canada. The data can be accessed through the Research Data Centres administered by Statistics Canada. The details and eligibility in obtaining data access can be found online (https://www.statcan.gc.ca/eng/rdc/process).

## Declaration

### Required statement for the analysis of Statistics Canada data at the Research Data Centre

The analysis presented in this paper was conducted at the Quebec Interuniversity

Centre for Social Statistics, which is part of the Canadian Research Data Centre Network (CRDCN). The services and activities provided by the QICSS were made possible by the financial or in-kind support of the Social Sciences and Humanities Research Council (SSHRC), the Canadian Institutes of Health Research (CIHR), the Canada Foundation for Innovation (CFI), Statistics Canada, the Fonds de recherche du Québec - Société et culture (FRQSC), the Fonds de recherche du Québec - Santé (FRQS) and the Quebec universities. The views expressed in this paper are those of the authors, and not necessarily those of the CRDCN or its partners[40].

### Ethics review

The CHMS was approved by the Statistics Canada ethics committee. The informed consent was obtained from all participants or, if participants are under 18, from a parent and/or legal guardian.[11, 12] This secondary data analysis was approved by the ethics review committee at the Centre Hospitalier de l’Université de Montréal.

## Acknowledgements

Not applicable.

## Author contribution

YSC conceptualized and designed this study, managed and analyzed data, and drafted the manuscript.

## Data Availability

The CHMS data are not publicly available. It is against the Statistics Act of Canada to release the CHMS data. The collection of the CHMS data has been approved by the ethics committee within the governments of Canada. The CHMS data have been deidentified and maintained by Statistics Canada. The data can be accessed through the Research Data Centres administered by Statistics Canada. The details and eligibility in obtaining data access can be found online (https://www.statcan.gc.ca/eng/rdc/process).

## Regulations

All methods were carried out in accordance with relevant guidelines and regulations.

